# AWARENESS OF ANTHRAX DISEASE AND THE KNOWLEDGE OF ITS TRANSMISSION AND SYMTOMS IDENTIFICATION: A CROSS SECTIONAL STUDY AMONG BUTCHERS IN ILE-IFE

**DOI:** 10.1101/2025.10.15.25338124

**Authors:** Sunday Charles Adeyemo, Eniola Dorcas Olabode, Folashade Yetunde Aremu, Sunday Olakunle Olanrewaju, Calistus Adewale Akinleye, Blessing Ele Idris, Israel Abiodun Rabiu, Oluwafemi Obehi Are-Daniel, James Ebunoluwa Atolagbe, Raheem Omotayo Olaposi

## Abstract

Anthrax is a zoonotic disease of public health significance as it has led to the morbidity and mortality of human and livestock. Anthrax is caused by *Bacillus anthracis* which causes contamination of soil and water. Due to the grazing nature of cattle, they are mainly affected by anthrax among other herbivores.

The study was a cross sectional study among 380 respondents selected using multistage sampling technique. Data was collected using a pretested, semi-structured interviewer-administered questionnaire. Data collected were analyzed with the use of IBM Statistical Package for Service Solutions (SPSS) version 25 software. Descriptive analysis was done for all variables.

Majority of the respondents (78.9%) were not aware about anthrax. More than half of the respondents (56.2%) agreed that anthrax can be transmitted through contaminated soil. Majority of the respondents (87.5%) who are aware of anthrax reported that there is relationship between anthrax and sudden death of animal. Majority (62.5%) said animals with anthrax die suddenly without illness and have dark un-clotted blood flow from their body openings. Majority (75.0%) said that carcass of animals with anthrax do not get stiff.

The study concluded that there is poor awareness about anthrax and its transmission among respondents. However, among respondents who were aware of anthrax, majority have good knowledge about identification of anthrax symptoms.

## BACKGROUND

Anthrax is a zoonotic disease of public health significance as it has led to the morbidity and mortality of human and livestock. It is also associated with economic loss due to reduced livestock trade. [1] It is seen in one of the three classical clinical forms; cutaneous, gastrointestinal and inhalation. Cutaneous anthrax is the most common form, accounting for up to 95% of all cases. Inhalation form produces the most severe human disease followed by the gastrointestinal form. However, all three forms have the potential to progress to a fatal systemic infection. [2]

Herbivores are more susceptible to anthrax than carnivores and omnivores and they are therefore agents by which B. anthracis spores spread and contaminate the environment due to their ability to carry B. anthracis spores in fomites to new areas.[3] Anthrax is caused by Bacillus Anthrax which causes contamination of soil and water, due to the grazing nature of cattle, they are mainly affected by anthrax among other herbivores.[4]

Anthrax can be transmitted to humans through contact with or consumption of infected animals. Transmission of the disease to humans mainly occurs through contact or consumption of infected animal carcasses and contaminated animal products. The incubation period of anthrax in humans varies depending on the form of anthrax. The gastrointestinal takes 2-5days while the cutaneous form takes 2-3days. The pulmonary form is usually delayed till up to 1-2 months and could also be within 15hours. In humans, depending on the route of entry, clinical symptoms are usually non-specific buy in livestock, the disease occurs as sudden death.[5] Symptoms in human includes, headache, fever, nausea, vomiting, abdominal pain, edema, and chest pain. Farmers, shepherds, butchers and occupations that deal with leather works and bone processing are prone to contacting anthrax especially those with cuts while handling infected animals. [6]

Osun State government has warned the public about buying and selling of infected animals and has put measures in place to curb the spread of the disease. The Director of Vetenary Services, in her sensitization speech enlightened the butchers in Osogbo that infected animals are usually found dead without any signs of illness with dark unclothed blood flowing from body openings like the nose, mouth, ear, and anal region of the affected animals, also the carcass does not become stiff after death. [7]

Though Anthrax is a rare disease in developed countries, it is becoming endemic in developing countries. Approximately 20,000 to 100000 cases are reported annually, however, 64 million livestock farmers are at risk.[8] Surveillance data in Uganda in 2018 revealed 186 reported human cases and 721 reported livestock deaths. [9] Outbreaks has also been confirmed in different regions of Kenya. [1] The first animal case of Anthrax in Nigeria was reported in July 2023 with 8 death of livestock who were observed to have been bleeding from external orifices without blood clotting. This was after the West Africa outbreak in Ghana in June 2023. The case was in a multi-specie animal farm comprising of cattle, sheep and goats located at Gajiri, along Abuja-Kaduna expressway Suleja LGA Niger State, where some of the animals had symptoms including oozing of blood from their body openings – anus, nose, eyes and ears. [10]

Since, the first outbreak of Anthrax has been reported, the National Center for Disease Control through the Executive director has raised an awareness about the possibility of infected animals being in other parts of the country, because prior to the confirmation of the first case of Anthrax in Niger state, there were movement of animals from the North down South during the Ileya festival, in which infected animals might have been transported. [11]

Osun State is notable for pastoral farming, and immigration of Fulani herdsmen. it also shares boundary with Kwara State, a North Central State with great population of Northerners. Osun State is also one of the states that received animals during the Ileya festival as many residents are Muslims. Ife, being one of the largest towns in Osun State will be studied.

Anthrax is a zoonotic disease; this means poor handling of an infected animal can cause a wide spread of anthrax in both animals and humans. Adequate knowledge on early recognition and identification of Anthrax symptoms in cattle is important in preventing the spread of the disease in the community. Hence, this study aims to assess the awareness of Anthrax disease and the identification of its transmission and symptoms among butchers in Ile-Ife, Osun State.

Farmers, shepherds, butchers and other occupations that deal with leather works and bone processing has been identified to be prone to contacting anthrax especially those with cuts while handling infected animals. [5] It has also been reported that there is a great dependence on livestock which makes people vulnerable to Anthrax among other zoonotic diseases. Livestock provide food, companionship, socio-cultural activities and are a source of income in various ways as they have an important economic role by sale and services of these animals and their products [12,13]

## METHODS

### a. Description of study area

The study location was Ile-Ife, a semi-urban area found in Osun - State, South-Western Nigeria. Ile-Ife administratively has two Local Government Areas in the town, Ife Central which has it’s headquarters at Ibadan Road and Ife East which has it’s headquarters in Oke-Ogbo. However, there are other local governments under Ife, located in other surrounding towns, they include Ife North which has its headquarters at Ipetumodu and Ife South which has its headquarters in Ifetedo and one Area Office located at Modakeke.

Ile-Ife and its environs occupy a large space in Osun State and accommodates a large number of immigrant pastoral farmers because of its vegetation. The dominant occupation in Ile-Ife is trading and farming, which encompasses a great number of livestock farmers and butchers.

### b. Study design

A descriptive cross-sectional study was used to assess knowledge at a given point in time among commercial butchers in Ile-Ife, Nigeria.

### Study population

This included commercial butchers in the four local government areas and the area office in Ife

### Inclusion criteria

Commercial butchers who have stalls and stands in main markets and surrounding areas in the four local government areas and the area office in Ile-Ife.

### Exclusion criteria

Commercial butchers who were sick at the point of data collection or unwilling to participate in the study.

### Sample Size Determination

Sample size will be calculated using Leslie Fischer’s Formula

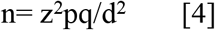

z = Standard Normal Deviate which is taken as 1.96 @ 95% confidence interval

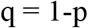

p= Expected Outcome which will be taken as 51.0% (0.51) which is the knowledge about human anthrax in Kisimu, Kenya (Mugo et al; 2019)

d= Precision which is taken as 5% (0.05)

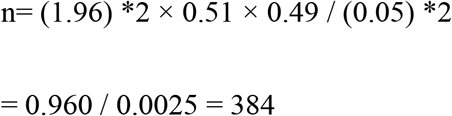

Adjusting for 10% non-response, an approximate of 450 respondents will be recruited.

### Data collection

A semi-structured questionnaire which was developed after review of relevant studies was administered to the respondents to assess their knowledge about Anthrax and their ability to identify its symptoms in animals. Data was collected from 12th to 17th February, 2024. A written informed consent was obtained from all participants.

### Sampling techniques

Two stage sampling technique was used to select the commercial butchers from the markets in Ile-Ife

**Stage 1**: The major markets in the Local Government Areas and Area Office was identified and two markets was randomly selected from each of the local government areas, making ten (10) markets.

**Stage 2**: Random sampling was used to select the commercial butchers in each market.

### Data Analysis

Data collected were sorted and analyzed with the use of IBM Statistical Package for Service Solutions (SPSS) version 25 software. Descriptive statistics was used to summarize the demographic characteristics and knowledge levels of respondents.

## RESULTS

Questionnaire was administered to 450 respondents, however, only 380 questionnaires were correctly filled. Giving a response rate of 84.4%.

Table 1 shows the demographic characteristics of respondents. More than half (53.9%) of the respondents were aged between 20-35 years. Majority of the respondents (71.1%) had secondary education. Two hundred and thirty (60.5%) of the respondents were Muslims while almost all respondents (92.1%) were from Yoruba tribe. About half (47.4%) of the respondents earn between 41000 – 100000 naira monthly. Majority of the respondents (88.2%) were male. Majority of the respondents (96.1%) rare cow and more than half of the respondents (69.7%) buy animal from the market and slaughter the animals themselves.

**Table 1:** Demographic characteristics of respondents (N=380)

| Demographic characteristics | Frequency | Percentage (%) |
| --- | --- | --- |
| **Age** |  |  |
| 20-35 | 205 | 53.9 |
| 36-45 | 125 | 32.9 |
| >45 | 50 | 13.2 |
| **Level of education** |  |  |
| Primary | 90 | 23.7 |
| Secondary | 270 | 71.1 |
| Tertiary | 20 | 5.3 |
| **Religion** |  |  |
| Christian | 150 | 39.5 |
| Muslim | 230 | 60.5 |
| **Tribe** |  |  |
| Yoruba | 350 | 92.1 |
| Hausa | 30 | 7.9 |
| **Income (in naira)** |  |  |
| 10000-20000 | 40 | 10.5 |
| 21000-40000 | 116 | 30.5 |
| 41000-100000 | 180 | 47.4 |
| >100000 | 44 | 11.6 |
| **Gender** |  |  |
| Male | 335 | 88.2 |
| Female | 45 | 11.8 |
| **Specie of live stock** |  |  |
| Cow | 365 | 96.1 |
| Goat | 10 | 2.6 |
| Pig | 5 | 1.3 |
| **Animal source** |  |  |
| Personal farm | 35 | 9.2 |
| Bought animal from market | 265 | 69.7 |
| Bought slaughtered animal from slaughter house | 80 | 21.1 |

Table 2 reveals the knowledge of respondents and their knowledge on the transmission of anthrax. Only 80 (21.1%) of the respondents have heard about Anthrax. The source of information for those who have heard about anthrax include friends/neighbours, Mass/social media and local market in which half of the respondents got the information from local market. (Figure 1)

**Table 2:** Knowledge about transmission of Anthrax.

| **Knowledge about transmission of Anthrax** | **Frequency** | **Percentage (%)** |
| --- | --- | --- |
| **Heard about Anthrax** |  |  |
| Yes | 80 | 21.1 |
| No | 300 | 78.9 |
| **Can anthrax be transmitted from animal to man (n=80)** |  |  |
| Yes | 15 | 18.8 |
| No | 65 | 81.2 |
| **Anthrax can be transmitted by handling infected animal (n=80)** |  |  |
| Yes | 30 | 37.5 |
| No | 50 | 62.5 |
| **Anthrax can be transmitted through contaminated soil (n=80)** |  |  |
| Yes | 45 | 56.2 |
| No | 35 | 43.8 |

**Figure 1:**
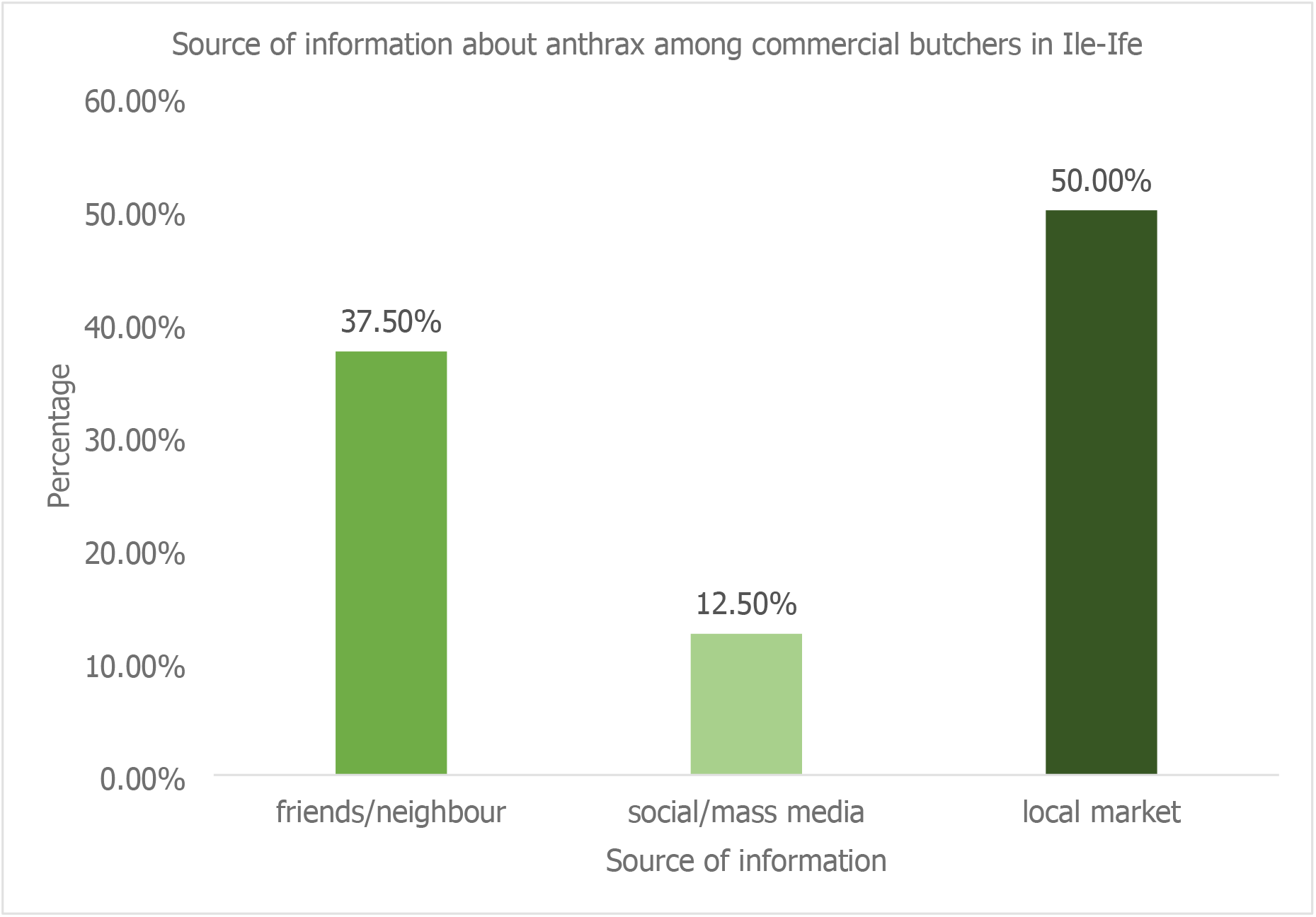
Source of information about anthrax among respondents

Out of the respondents who have heard about anthrax, only 18.8% agreed that anthrax can be transmitted from animal to man. Less than half (37.5%) agreed that anthrax can be transmitted by handling infected animal. However, more than half of the respondents (56.2%) agreed that anthrax can be transmitted through contaminated soil.

Table 3 shows the identification of anthrax among respondents. Majority of the respondents (87.5%) who are aware of anthrax reported that there is relationship between anthrax and sudden death of animal. Majority (62.5%) said animals with anthrax die suddenly without illness and have dark un-clotted blood flow from their body openings. Majority (75.0%) said that carcass of animals with anthrax do not get stiff.

**Table 3:** Knowledge about Identification of Anthrax symptoms.

| Symptoms of Anthrax (n=80) | Frequency | Percentage (%) |
| --- | --- | --- |
| **Relationship between anthrax and sudden death of animal** |  |  |
| Yes | 70 | 87.5 |
| No | 10 | 12.5 |
| **Animal dies suddenly without illness sign** |  |  |
| Yes | 30 | 37.5 |
| No | 50 | 62.5 |
| **Dark un-clotted blood flow** |  |  |
| Yes | 30 | 37.5 |
| No | 50 | 62.5 |
| **Carcass does not become stiff** |  |  |
| Yes | 60 | 75.0 |
| No | 20 | 25.0 |

## DISCUSSION

The study assessed the awareness of anthrax among respondents. The study also assessed the knowledge of transmission of anthrax and the identification of anthrax symptoms among respondents.

Awareness about anthrax was poor among respondents. Only a little above one-fifth of the respondents have heard about anthrax before the time of data collection. Half of the respondents who have heard about anthrax 4got the information from the local market. This is quite surprising as there was mass media announcement on anthrax by the state government. However, this poor knowledge can be attributed to some socio-demographic factors among respondents. Majority of the respondents had secondary education which could be a barrier in comprehending information. They might not have paid attention to the news. This is lower than the level of awareness about anthrax disease found in the study of Cadmus et al; (2024) who reported an awareness level of 71.2% among 7 selected states in Nigeria.

Knowledge about transmission of anthrax was poor among respondents. Out of the respondents who have heard about anthrax, less than one-fifth agreed that anthrax can be transmitted from animal to man. About two-fifth agreed that anthrax can be transmitted by handling infected animal. However, about 6 out of 10 agreed that anthrax can be transmitted through contaminated soil. This low level of knowledge can be attributed to the fact that 7 out of 10 respondents buy animals from the market, this means they will most likely buy healthy animals and are not aware about any disease that could attack animals.

Majority of the respondents who were aware of anthrax had good knowledge about the symptoms of anthrax. About 9 out of 10 respondents reported that there is relationship between anthrax and sudden death of animal. A little above 6 out of 10 respondents said animals with anthrax die suddenly without illness and have dark un-clotted blood flow from their body openings. More than 7 out of 10 respondents said that carcass of animals with anthrax do not get stiff. This is similar to the findings in the study of Cadmus et al; (2024) who reported bleeding from natural openings, dark un-clotted blood and sudden death has symptoms of anthrax identified by respondents.[14]

## LIMITATION OF THE STUDY

Being a cross-sectional study, the study has a limitation of generalisability. Also, the study was carried out in an area with low-level of literacy, which could have had effect on the results of this study.

## CONCLUSION

The study concluded that there is poor awareness about anthrax and its transmission among respondents. However, among respondents who were aware of anthrax, majority have good knowledge about identification of anthrax symptoms. The poor knowledge on the transmission of anthrax disease can lead to poor attitude to the prevention of anthrax disease among animals and consequently the spread of Anthrax disease among humans and animals.

Awareness about anthrax should increase using various means of communication which include mass media, posters and community awareness.

## Ethical Approval and Consent to participate

This study was approved by the Research Ethics Committee of the Osun State University, Osogbo (UNIOSUNHREC 2024/002).This study was conducted according to the Declaration of Helsinki for Medical Research involving Human Subjects.

### Right of decline/withdraw from study

Respondents were told that participation is voluntary and they will not suffer any consequences if they chose not to participate.

### Confidentiality of data

All information gathered were kept confidential and participants were identified using serial numbers.

### Consent form

A written informed consent was obtained from all participants.

### Non-maleficence

No harm is intended nor befell any respondent in the course of the research study. Respondents were reassured of this.

## Acknowledgements

The authors wish to acknowledge the respondents and the spouses of the authors for their understanding during the course of this study.

## Data availability

The data for this study has been presented in the manuscript

## Author’s contributions

Sunday Charles Adeyemo-Study design, data analysis and manuscript writing; Eniola Dorcas Oluwafemi - Study design, data analysis and manuscript writing; Folashade Aremu-Data collection; Sunday Olanrewaju-Supervision; Calistus Akinleye-Supervision; Blessing Ele Idris-Data collection; Isreal Abiodun Rabiu-Data collection; Obehi Are-Daniel -Graphics; James E. Atolagbe-Supervision, Raheem Omotayo Olaposi - Data collection.

